# Health Systems Reforms in Bangladesh: An Analysis of the Last Three Decades

**DOI:** 10.1101/2023.10.11.23296847

**Authors:** Treasure Udechukwu, Thierno Oumar Fofana, Louise Carnapete, Shams Shabab Haider, Suhi Hanif, Lucie Clech, Valéry Ridde

## Abstract

**Objective:** We reviewed the evidence regarding the health sector reforms implemented in Bangladesh within the past 30 years to understand their impact on the health system and healthcare outcomes.

**Method:** We completed a scoping review of the most recent and relevant publications on health system reforms in Bangladesh from 1990 through 2023. Studies were included if they identified health sector reforms implemented in the last 30 years in Bangladesh, if they focused on health sector reforms impacting health system dimensions, if they were published between 1991 and 2023 in English or French and were full-text peer-reviewed articles, literature reviews, book chapters, grey literature, or reports.

**Results:** Twenty-four studies met the inclusion criteria. The primary health sector reform shifted from a project-based approach to financing the health sector to a sector-wide approach. Studies found that implementing reform initiatives such as expanding community clinics and a voucher scheme improved healthcare access, especially for rural districts. Despite government efforts, there is a significant shortage of formally qualified health professionals, especially nurses and technologists, low public financing, a relatively high percentage of out-of-pocket payments, and significant barriers to healthcare access.

**Conclusion:** Evidence suggests that health sector reforms implemented within the last 30 years had a limited impact on health systems. More emphasis should be placed on addressing critical issues such as human resources management and health financing, which may contribute to capacity building to cope with emerging threats, such as climate change.

## INTRODUCTION

Over the past fifty years, several health sector reform initiatives have been implemented in South Asia. In Bangladesh, these reform initiatives have enabled the country to achieve development goals, such as reducing under-five mortality rates from 125 to 27 per 1000 live births between 1995 and 2019. Over the same period, the proportion of births attended by skilled health personnel rose from 9.5% to 58%, and overall life expectancy at birth from 47 to 74 years (1). Despite these strides, there needs to be more exploration regarding the strengths and weaknesses of both past and current health policies. To our knowledge, there have been no comprehensive literature review on the impact of health sector reforms in Bangladesh. Health sector reforms can be a powerful tool for countries to adapt their healthcare system in response to demands and external shocks (2,3). As Bangladesh is ranked seventh on the Global Climate Risk Index 2000–2019 (2), there is an increasing need to understand the effectiveness of health policies and reforms in Bangladesh in preparation for tackling emerging climate and climate-associated issues (4), including the recent COVID-19 pandemic (5).

The Bangladesh health system is pluralistic and consists of the government, the private sector, the non-governmental organisations (NGOs) and the international donor organisations. Two ministries within the government are responsible for providing and regulating healthcare. While the Ministry of Health and Family Welfare (MoHFW: oversees both public and private sector) provides public sector healthcare up to the tertiary level in rural and urban areas, the Ministry of Local Government, Rural Development and Cooperatives is responsible for primary healthcare in urban areas (urban primary healthcare is the responsibility of city government/municipality) (6). The MoHFW has two branches: the Directorate General of Health Services and the Directorate General of Family Planning. With regard to structures, 53 District Hospitals, 425 Upazila Health Complexes, 1469 Union Health and Family Welfare Centres, and 12 248 community clinics (CC) at the ward level were managed through these two branches of the ministry in 2015 (6). However, the two ministries’ division of responsibilities within the health system is unequal: the MoHFW is in charge of health system planning at the central level, with little involvement from the Ministry of Local Government, Rural Development, and Cooperatives. This implies that the local government needs more control over how the local health system is designed. But the ministry of local government, despite the name is also a part of the central government. More involvement by this ministry does not necessarily equal more involvement by local (non-central) government. Urban primary healthcare (PHC) is left out of nationwide planning related to healthcare, which is done by the MoHFW.

Additionally, the health system relies heavily on the private sector (formal and informal) and NGOs to fill gaps in service delivery (6). The NGO sector has been emerging as an increasingly growing sector. In 1997, NGOs sourced 6% of total health expenditure, which increased to 9% in 2007. While the government, private sector, and non-governmental organisations focus on service delivery, financing, and healthcare staffing, donors are critical in financing and designing the government’s health programs. This dependency on donor organisations limits the government’s involvement in planning context-specific health policies (5). This can be explained through the lack of “country ownership” in designing and implementing donor-supported initiatives, a common issue in global health (7). In simpler terms, this refers to the alienation of local authorities familiar with the health policy context from the actual decision-making process of donor organisations.

We aim to understand the impact of health system reforms on the health system of Bangladesh.

## MATERIALS AND METHODS

We used the scoping review method developed by Arksey and O’Malley (8). We followed the Preferred Reporting Items for Scoping Reviews (PRISMA-ScR) guideline (9), see PRISMA-ScR-Checklist as Supporting information 1. This method was appropriate because it gave us an overview of the available literature in this specific field (10). A first version of the protocol has been published (11).

### Research questions

Our research question was: what are the impacts of health reforms in Bangladesh on health systems? Our specific objectives were: (1) identifying the health sector reforms implemented in Bangladesh within the past 30 years and (2) understanding the impact of those reforms on the health system.

### Search strategy

We carried out a systematic literature search to explore the subject in depth. The methodological approach followed key steps to ensure exhaustive coverage of relevant sources. We targeted databases in health sciences research, including PubMed, Web of Science, SCOPUS, and Google Scholar. Our search was conducted from March 2021 to April 2021, with a new search for March 2023 and June 2023 to check recent publications. The search strategy was designed to be comprehensive and precise. We used the broad search terms ‘reforms’ AND ‘health systems AND ‘Bangladesh’ to ensure we captured a wide range of articles. In addition, we incorporated controlled vocabulary terms such as ‘healthcare system’, ‘health policy’ and ‘healthcare sector’ to further refine our search. The Population/Concept/Context framework recommended by the Joanna Briggs Institute guided our research in identifying key research elements, ensuring consideration of the population of interest, the relevant concepts, and the specific context of Bangladesh. The final detailed and elaborated research strategy is in the supplementary materials as Supporting information 2.

### Selection process

Search results were imported into Rayyan, the online systematic review software. Two reviewers (TU and LC) independently screened titles and abstracts of identified papers and then again screened the full text. Retrieved studies were evaluated according to the inclusion criteria and focused on the topic of interest. Discrepancies in reviewers’ responses at abstract and full article screenings were resolved through discussion.

### Inclusion and Exclusion Criteria

Articles were included if they: (1) identified health sector reforms implemented in the last 30 years in Bangladesh; (2) focused on health sector reforms impacting health system dimensions; (3) were published between 1991 and 2023 in English; (4) were full-text peer-reviewed articles, literature reviews, book chapters, grey literature, or reports. Additionally, two studies were included from the references of studies that were selected during full-text screening. Health sector reforms were defined as ‘sustained, purposeful changes to improve the efficiency, equity, and effectiveness of the health sector’ (12). Health sector reforms focus on setting policy objectives covering core functions of the health system, revising policies, and reforming the institutions that implement these policies (13).

### Charting the data

We used an Excel spreadsheet to extract all relevant data such as author(s), title, year of publication, journal name, methodology, objectives, key findings, reforms, and impact on health system dimensions and access to care. We extracted data from the included articles based on the policy analysis framework (process, content, actors, context) and the impact on the healthcare system (building blocks) and access to care (availability and affordability) (14–17). Quality assessment of the studies (MMAT) has been used to describe the methods of each paper (18).

Data extraction was initially conducted by two reviewers independently (TU, LC), and verified by a third (VR or LC). Differences between reviewers were resolved by reaching a consensus. The data extraction form is provided in the Supplementary Materials. Finally, a narrative synthesis (19) was conducted, and the findings were tabulated to summarise their characteristics and enable comparison and analysis across studies. The key results of all included studies are reported in Supporting Information 3.

### Ethics approval, consent to participate, competing Interests

This is a scoping review with no empirical data collection, so no ethical approval is required. However, this review is part of a research project that has obtained an Ethics approval from the Institutional Review Board (IRB) of the BRAC James P Grant School of Public Health, BRAC University (ref: IRB-19 November 20–050) in Bangladesh. The authors have declared that no competing interests exist. See S4 Questionnaire for inclusivity for global health research process.

## RESULTS

### Search findings

Our search yielded 3559 studies, 3413 from scientific databases and 146 from grey literature databases) from which we excluded 1029 duplicates. Title and abstract screening was conducted on the remaining 2530 studies. After this initial screening, 2492 studies were excluded due to the following reasons: (1) published before 1991, (2) no keywords (reform or policy), (3) not focused on reforms in the context of the health system. Full-text screening was conducted on the remaining 38 studies. Three studies were excluded since their full-text versions were unavailable, and another fifteen were excluded since they either discussed policy reforms before 1991 or had findings irrelevant to our research question. An additional three studies were included. Three after screening the references of our included studies. This resulted in a final list of 24 studies that met our eligibility criteria (Appendix 2). Figure 1 shows the PRISMA diagram of the study selection process.

**Figure 1:**
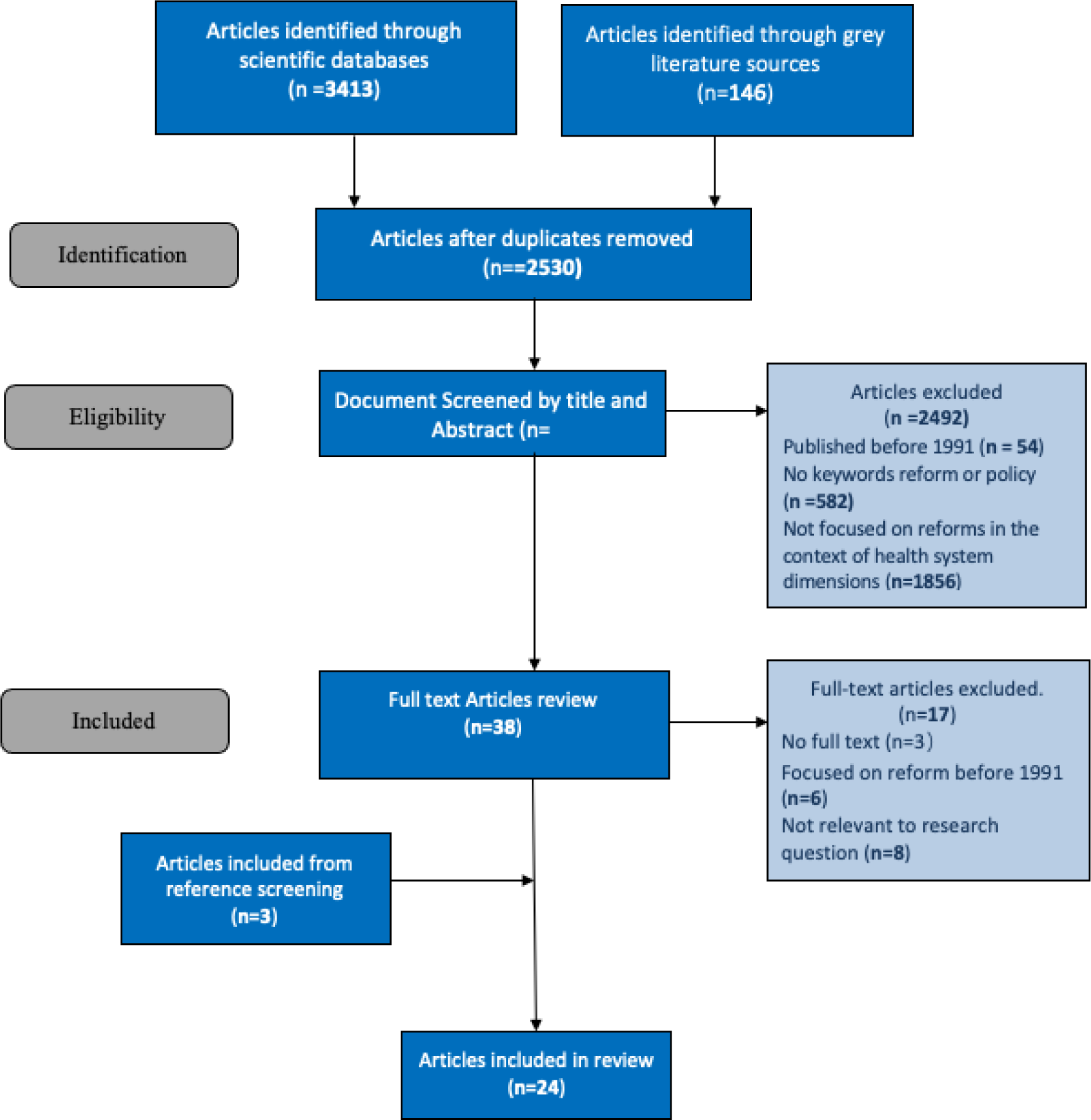
PRISMA Flowchart.

### Study characteristics

The publication year of the included articles ranged from 2000 to 2022, with the most representation from the latter half (Figure 2). The first authors in eleven studies were affiliated with institutions in Bangladesh (20–30), nine studies were affiliated with both institutions inside and outside of Bangladesh (31–39) and four studies were affiliated with foreign institutions (40–42). We identified only four articles that adopted a conceptual framework to guide their work: two by the same principal author used the Health Policy Triangle framework (26,28), one used the Universal Health Coverage (UHC) Cube (23), and one developed a new conceptual framework (31).

**Figure 2:**
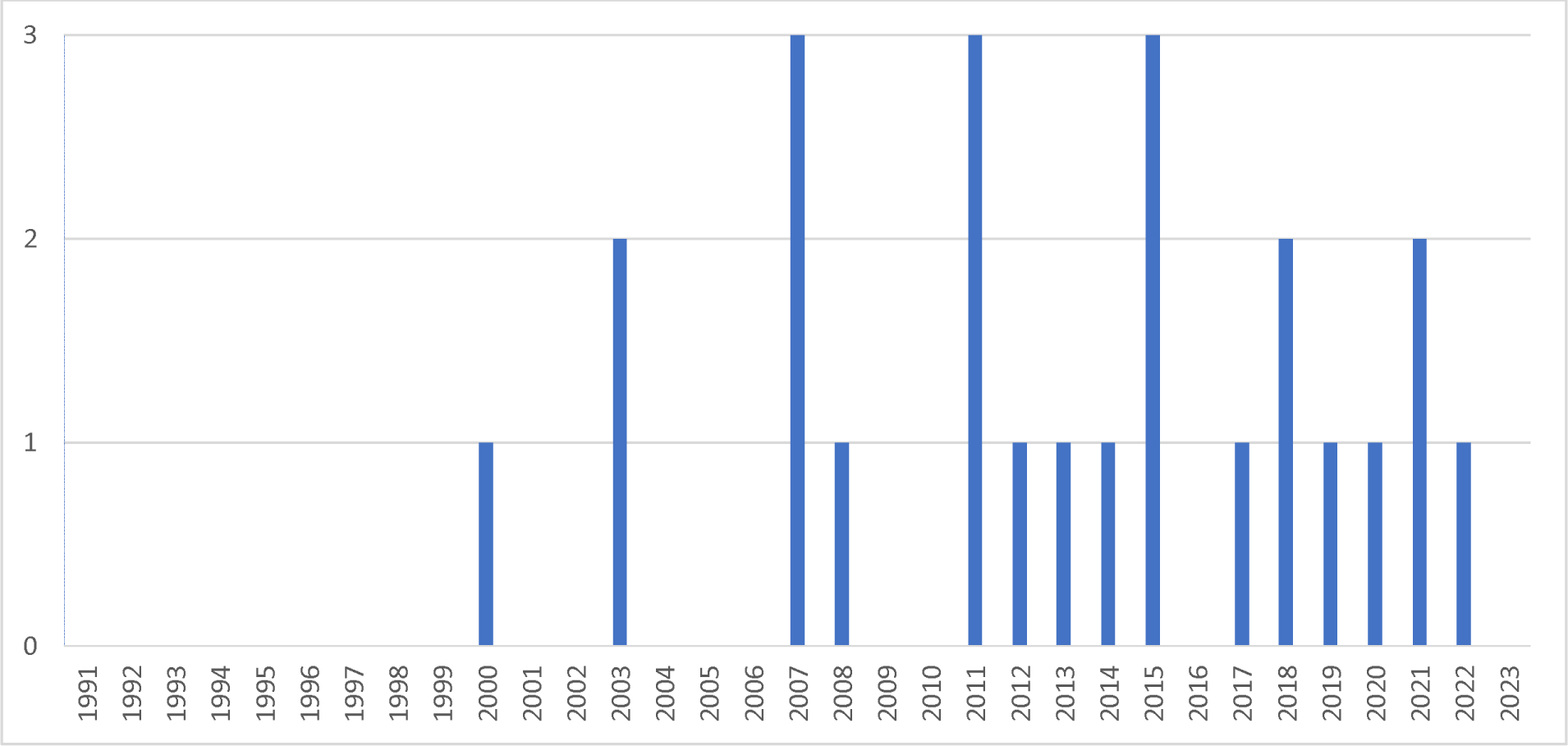
Number of included articles by publication year (n=24)

Eleven studies were review articles and reports that could not be evaluated using the MMAT tool (Figure 3). The selected studies included mixed methods (n=3), qualitative (n=6) and quantitative studies (n=4). Five studies did not clearly state their objectives (28,34–36,40). There is no information or clarity on properly considering the researcher’s influence on the research for five qualitative studies. Three studies were rated with all indicators positive (yes) (23,31,32), while the remaining ten studies were rated with at least one indicator positive. The indicator was labelled ‘can’t tell’ when information was missing or unclear descriptions.

**Figure 3:**
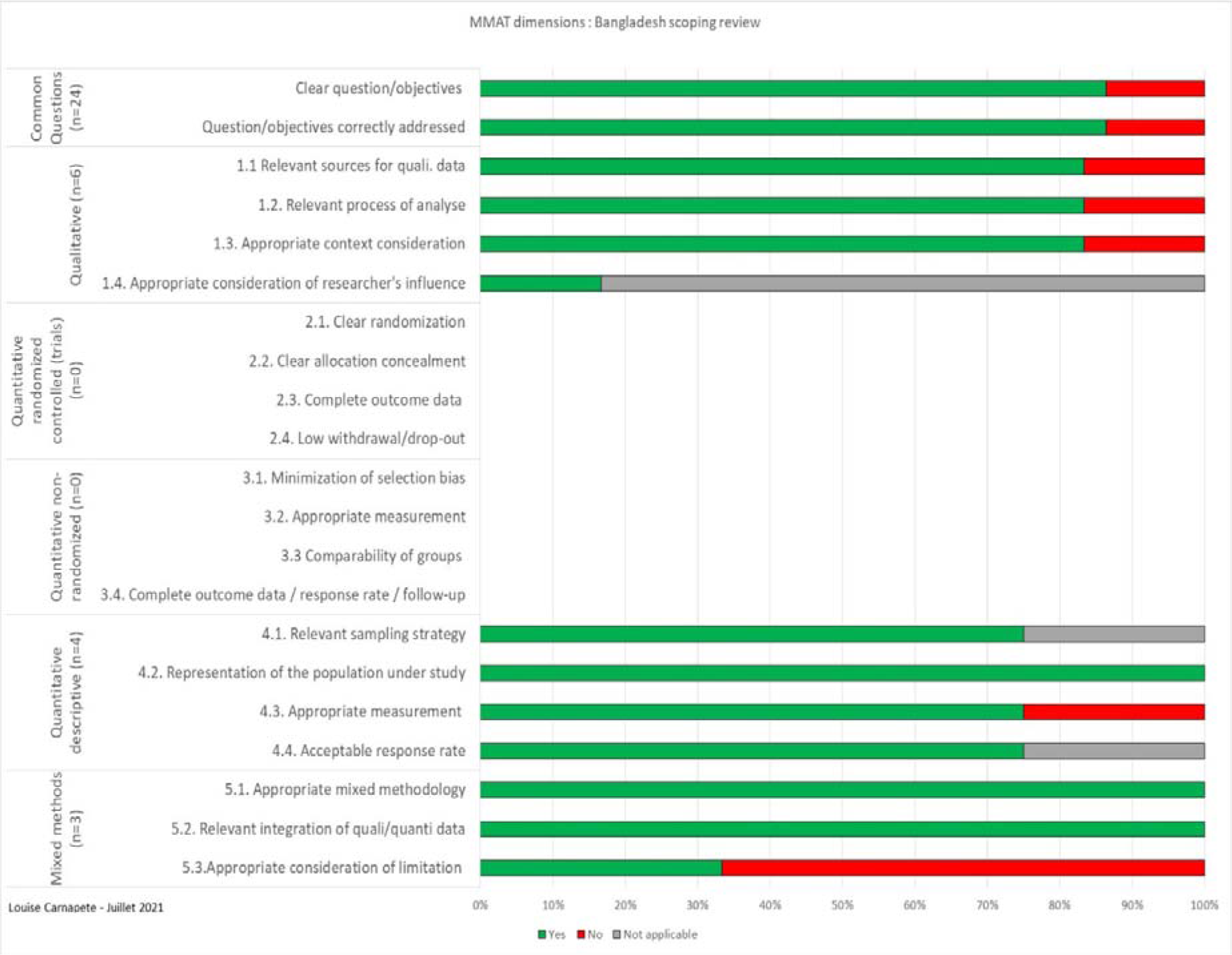
Quality assessment using the Mixed Method Appraisal Tool (MMAT), n = 13.

### Health Sector Reforms in Bangladesh

We present our findings through the lens of the policy analysis framework. Figure 4 presents a summary history of the reforms identified in the selected articles.

**Figure 4:**
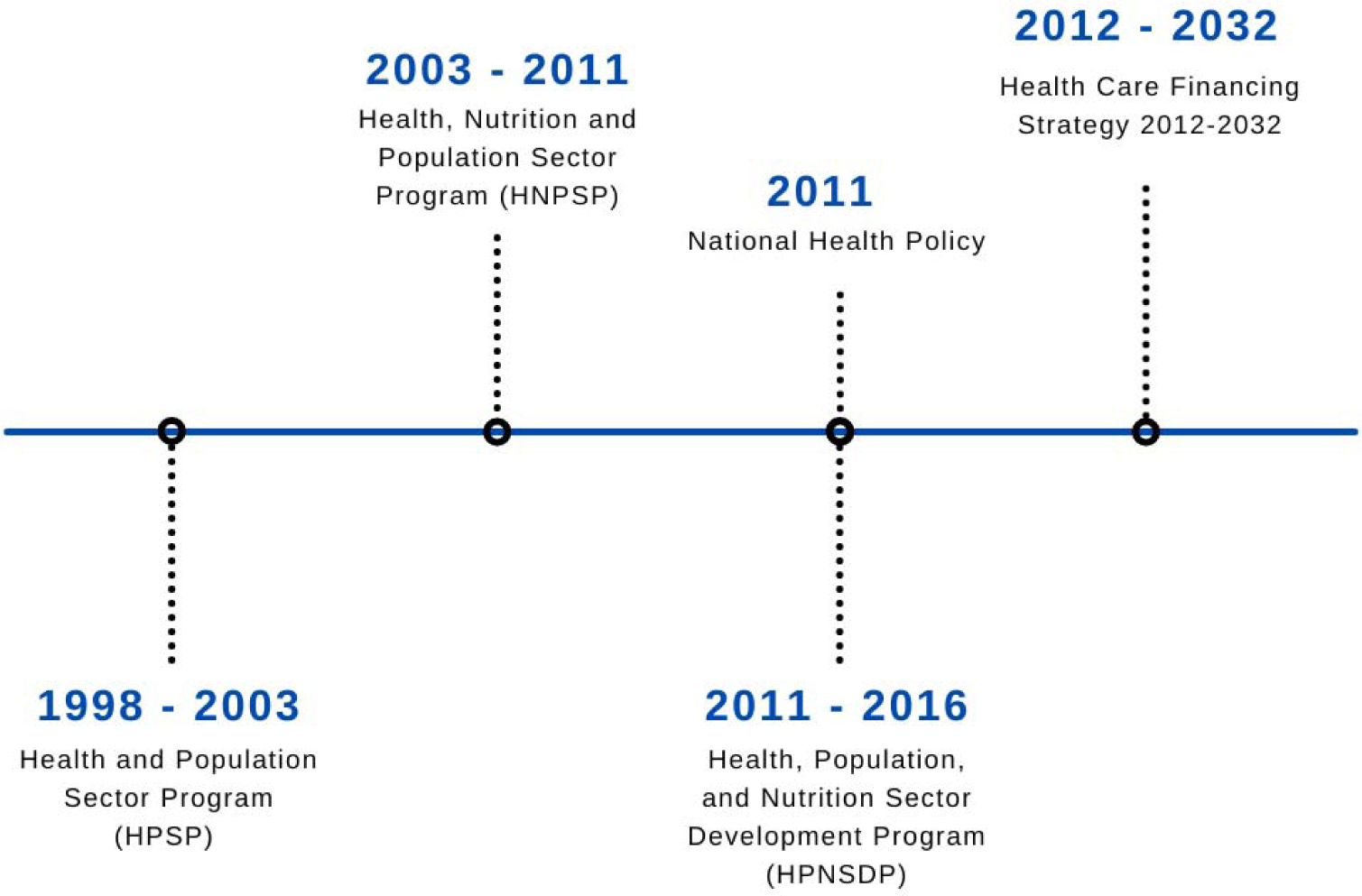
Health Systems Reforms in Bangladesh Timetable.

### Context

After independence in 1971, Bangladesh struggled with high mortality rates, poor female literacy, vulnerability to natural disasters, and starvation (31). During this period, health sector operations were primarily focused on population control and primary healthcare for the poor and disadvantaged (20). To achieve this goal, development partners (DPs) played a crucial role in financing a series of family planning and health projects (36,42). However, these projects could have been more effective and sustainable (42).

In 1990, Bangladesh’s then-military regime attempted to implement the first National Health Policy (NHP), strongly influenced by the Health and Population Sector Strategy (HPSS) (20). The 1982 National Drug Policy, successfully adopted about eight years earlier in the same regime, was one of the critical drivers for the planned 1990 NHP (22). The NHP resulted from recommendations by the Health Care System Improvement Committee, formed in 1987 (22). However, the military regime failed to adopt the new NHP. This failure was due to fierce resistance from internal players such as political parties (i.e. pre-democratic), private and business associations and the considerable influence of external actors, notably multinational companies and international NGOs, who lobbied intensively for their interests and perspective. In 2000, the Government of Bangladesh (GoB) adopted a new NHP, with an overall framework encompassing 15 distinct objectives, guided by ten fundamental policy principles, and supported by 32 strategic initiatives. This new National Health Policy was met with resistance from the post-military dictatorship government and never saw the light of day (43). These failures led the Bangladeshi health system to operate without a National Health Policy until 2011 (22).

Around the 1990s, significant bilateral and multilateral donor institutions adopted the Sector Wide Approach (SWAp), designed to provide aid to developing countries via a single channel (20,36). At this time, Structural Adjustment Programs had been adopted by Bangladesh as a condition for funding from the International Monetary Fund (IMF) and the World Bank. It began with ‘structural and sectoral adjustment loans’ from the World Bank in 1980 (37). One-third of the overall international development aid commitments to Bangladesh between 1980 and 1996 were Structural Adjustment lending that amounted to USD 5.97 billion, involving 93 projects, among which USD 4.65 billion was disbursed (37). At this time, economic liberalisation and privatisation reduced government funding for health services, impacting the nature of health service delivery and increasing the influence of donors on public health policy (37).

Bangladesh’s dependence on foreign financing to implement public policy made it necessary to adhere to the agenda of the international donor community (20). In 1996, the GoB worked towards a comprehensive sector-wide strategy that included a structural and organisational reform agenda by the MoHFW further to obtain loans from its DPs (33,38). The Health and Population Sector Strategy (HPSS) implemented in 1997 resulted from this work (37,42,43). This strategy supported several institutional and governance reforms that enabled the health sector to move towards increased efficiency and cost-effectiveness (20).

## Content

### Objectives of the reforms

The MoHFW envisaged that SWAp would address the lack of an integrated, cost-effective health system in three ways: (1) better coverage of essential health and family planning services would be guaranteed through technical assistance and coordinated financing; (2) cost-effectiveness of service delivery through leveraged sector reforms; and (3) participation by NGOs and the private sector in service delivery (42).

The Health and Population Sector Program (HPSP) was the inaugural SWAp, from 1998 to 2003 (20,32,35,42,42). It was led by the government and funded by pooled and bilateral funding from the government and DPs. The HPSP’s primary goal was to use a “one-stop” service model to decentralise the delivery of the essential service package of PHC to rural communities using CCs (35,42).

In 2003, the second SWAp program, the Bangladesh Health, Nutrition and Population Sector Program (HNPSP), was created. HNPSP ran between 2003-2011, aiming to improve the availability and use of user-oriented, efficient, fair, affordable and accessible health, nutrition, and population quality services (20,34,42).

The third SWAp program, the Health, Population, and Nutrition Sector Development Program (HPNSDP), was implemented between 2011–2016. The primary goal was strengthening health systems and improving health and family planning services. The overarching goal of all three SWAps had been to improve the access and utilisation of an essential package of health, population, and nutrition services, particularly among poor women and children (42).

In 2011, the GoB implemented a new and comprehensive health policy. This policy paper mostly aligned with the Millennium Development Goals, such as reducing maternal and infant mortality rates (22). This updated health policy aimed to ensure accessibility to PHC and emergency services for all; to extend the coverage of quality, equity-based services for all; and to increase community demand for healthcare while respecting rights and dignity (22).

Under the HPNSDP, a key policy document addressing UHC in Bangladesh titled ‘Health Care Financing Strategy 2012-2032: Expanding Social Protection for Health towards Universal Coverage’, was produced by MoHFW’s Health Economics Unit (HEU). This document, which was designed to address health funding issues for the next 20 years, also proposed ways to combine funds from tax-based budgets with proposed social health protection schemes (including for the poor and the formal sector), existing community-based schemes, additional advance payment schemes, and donor funding for all segments of the population, starting with the poorest (23).

### Planned Reform Initiatives

During 1998–2016, several reform initiatives were planned under the health sector reform programs to improve the health system and service delivery.

- A change in management from a project-based approach to a sector-wide approach.
- Decentralization in the delivery of the essential service packages utilizing the concept of a ‘one-stop’ service model to provide essential health and family planning services to rural communities from static Community clinics.
- Diversification of services through the involvement of stakeholders, including NGOs, and the implementation of diverse policies and initiatives (e.g., gender strategy, drug policy, etc.).
- Improving financial management and procurement processes, outsourcing services, and establishing an NGO contracting system.
- Developing a national healthcare financing framework and expanding the Demand Side Financing program based on its evaluation.
- Creating a Monitoring and Evaluation strategy and implementing an online procurement tracking system.
- Developing cadres of health assistants and family welfare assistants, as well as training personnel and health personnel.
- Creating a Human Resource (HR) plan, establishing a functional HR Information System, and implementing incentive packages to deploy and retain a critical health workforce in remote and rural areas.
- Addressing the challenge of skilled-birth attendance by training community-based skilled birth attendants and/or nurse-midwives, midwives, and family welfare visitors, and streamlining the recruitment and promotion of nurses.
- Construction of many community clinics.

#### Process

The HPSS of 1997, developed collaboratively by the GoB and DPs, signalled the decision to shift from a project-based modality to SWAp. The GoB agreed to carry out the health sector programs through operational plans supervised by a Line Director. Each operational plan contained a set of SWAp tasks, funds, and periodic performance and resource reviews headed by the MoHFW. An annual program review is conducted by the GoB, based on performance indicators agreed upon by the MoHFW and DPs (42).

However, there were changes to the implementation process during the HPNSDP (2011–2016) based on learnings from the previous SWAp programs. Additionally, the partnership agreement between the GoB and its DPs for the program was strengthened further through a joint financing agreement.

The MoHFW shared the Annual Development Program budget with the DPs to ensure that priority interventions were sufficiently resourced. The Annual Progress Report was synced with the preparation of the operational plans and annual development program by the MoHFW and DPs to ensure that the yearly progress report’s recommended actions were included and implemented. The HPNSDP had a coherent multi-year integrated and consolidated technical assistance strategy, which was prepared to assist implementation and enhance the MoHFW’s institutional capacity at various levels and improve the focus on achieving outcomes and carrying out agreed-upon reforms (40).

### Limiting and Supporting factors

In the early years of SWAp, the MoHFW’s ownership and leadership was relatively weak, as evidenced by the DPs’ reluctance to give up control over aid management and coordination due to ‘weak government capacity, inadequate accountability and compromised integrity’. Several reform projects failed during the first two SWAp programs, primarily due to poor adaptation of planned activities to changing policy environments. The military dictatorship attempted to draft a NHP but failed due to heavy internal and external stakeholder pressure (i.e., political parties, NGOs, etc.).

The HPSP was not fully implemented as intended. The World Bank Implementation, Completion and Results report rated the HPSP as ‘unsatisfactory’. One of the main issues reported was the unification of health and family planning services. The unification was resisted at the district levels and above but supported at the local level (20,32). In 2001, a change in government halted the unification, which was later reversed in 2003. The new government also halted the construction of CCs after 2001 (32). Findings show that the unification of health and family planning wings, withdrawal of domiciliary services, and inefficient management, including insufficient supply of drugs and poor supervision, were the main barriers to achieving the expected results during HPSP (20). Reform planners ignored the role of context in their reform objective planning and expected that the workforce would be a passive element in reform implementation (33)

A quarter of the funding administered by the World Bank was allocated under HNPSP for performance-based financing (PBF) between 2003 - 2011. This leveraged changes/reforms that contributed to SWAp objectives and promoted the achievement of key health outputs (42). This modality was unsuccessful, and the amount set aside for PBF in the first few years was not disbursed, owing to insufficient incentives for results and weak links between the agency responsible for meeting the targets and the recipient of the PBF funds. Moreover, some DPs exited the Health, Nutrition and Population sector (e.g., AusAID, EU, The Netherlands, GIZ) (42).

Despite several limiting factors, the GoB has remained committed to health improvements. Bangladesh has a favourable political climate that supports health sector reforms aided by institutional continuity of civil servants and partnerships between the governmental and non-governmental sectors (20,31).

#### Actors

Under SWAp in Bangladesh, the GoB plays a significant role alongside the World Bank, IMF, DPs, and other stakeholders, such as health professionals, traditional practitioners, and non-profit health workers (24,31,37). Over the past 15 years, the GoB’s financial contribution to SWAp programs has increased and the share from DPs has gradually decreased. The GoB contributed USD 2.2 billion (62%) to the HPSP, USD 5.4 billion (67%) to the HNPSP, and USD 7.7 billion (76%) to the HPNSDP (42). The remaining shares of all three programs were co-financed by the World Bank, Canada, Germany, The Netherlands, Sweden, the United Kingdom, the European Union, UNFPA, Australia, and the United States (42).

Although the GoB contributes financially to certain programs, such as SWAPs, it is undeniable that donors exert a significant influence on the health policy-making process. The World Bank’s influence in Bangladesh’s healthcare sector is a good example. It not only provides fundings for health programs, but also plays a major role in setting priorities for the allocation and management of financial resources (37). The MoHFW is the primary health policy-making body in Bangladesh and in charge of capacity building (28). The change in political power in 2009 influenced the MoHFW to adopt a new NHP in 2011 (43). The Health Care Financing Strategy 2012-2032 developed by the HEU was actively supported by donor partners (23).

## Impact on Health System Dimensions

### Health Sector Management

Three studies identified decentralisation of the administrative systems following the introduction of health sector reforms (22,24,29). Decentralization in Bangladesh happened in power de-concentration, with some authority and responsibility transferred from the central level to senior program managers, often known as Line Directors, who have complete autonomy in administrative and financial affairs.

The administration in the health sector was also decentralized at the sub-district and lower levels to increase the public health sector’s capacity to deliver high-quality services (22,24,29). However, actual decentralization under the health sector reform programs had yet to be achieved as the healthcare system never undertook a devolution process [26]. The operations and power had been decentralised to senior program managers placed at the national level rather than to lower tiers of administration such as district, sub-district, and union level (22,24,29). This impacted the allocation of health resources at the local levels and restricted the local authorities from addressing local issues (24,29).

On the other hand, one of the reform initiatives planned under the health sector reform programs was diversification of services through the involvement of other stakeholders. One study reported that under the HNPSP, diversification of services was achieved through official collaborations between the private and public sector. This partnership resulted in a shift of government’s role from ‘provider’ of health services to that of a ‘purchaser’ (20).

The health sector reform introduced the unification of the MoHFW’s health and family planning wings. But this reform resulted in conflicts as national and district-level family planning directors lost authority to lower-level personnel (33). The unification of health and family planning wings was reversed along with the restoration of domiciliary services (20).

### Workforce

Four studies reported improvements in the availability of qualified health professionals and shortages with the introduction of the health sector reforms (21,27,36,42). Under the health sector reform programs, the density (per 10,000 populations) of physicians and nurses has increased over the last decade (from 1.9 physicians and 1.1 nurses in 1998 to 5.4 physicians and 2.1 nurses in 2007). The density of dentists also increased but is still quite low (from 0.01 in 1998 to 0.3 in 2007) (21). This increase in qualified health professionals explains the significant growth in outpatient consultations and admissions in government health institutions (42).

Despite these improvements, physician retention in rural areas and a skewed health workforce toward physicians remains a severe problem (36,42). In addition to this imbalance, absenteeism is one of the biggest challenges facing the healthcare system regarding HR for health. This challenge is exacerbated by structural factors such as lack of effective regulation, housing problems, insecurity and precarious working conditions (30,39). The density of formally qualified healthcare professionals (physicians, nurses, and dentists) is relatively low at 7.7 compared to other South Asia countries. The nurse-to-doctor ratio of 0.4 (2.5 times more doctors than nurses) falls significantly short of the international standard of three nurses per doctor (21). Based on the doctor-to-population ratio in low-income countries, Bangladesh has an estimated shortfall of about 60,000 doctors, 280,000 nurses, and 483,000 health technologists. The overwhelming urban bias in the distribution of qualified healthcare professionals observed over a decade ago remains a persistent problem. Many of these health providers are disproportionately concentrated in Dhaka and Chittagong (21,27). This shortage and maldistribution of health care professionals, support and technical workers negatively affects the commitment to attain health reform objectives (36).

### Finance

Five studies cited increased finance for health services with the introduction of health sector reforms. It also reports on poor financial management, low public funding and high out-of-pocket payments as significant problems of the Bangladesh healthcare system (20,22,27,29,42). Under the HPNSDP, DPs financing grew from around USD 800 million to USD 1.8 billion, enabling the MoHFW to plan and implement key health reform initiatives. DP financing also explains the increase in the MoHFW budget execution capacity from spending 76% of its annual allotment in 2004–2005 to 89% in 2011–2014 (42). The introduction of essential service packages during the HPSP period led to more spending on PHC (between 60% and 70% of public expenditure), focused attention on maternal and child care, and shifted the attention from hospitals to PHC services used by the lower income communities (20). This meant improved access to basic essential services for those in the lowest economic brackets, mostly in rural areas.

However, the health sector reforms have not impacted increasing public health expenditure and providing financial protection. Two studies reported that Bangladesh spent approximately 3.4% of its GDP on health in 2014, with the government contributing about 1.1% (22,29). The total health expenditure was approximately USD 12 per capita per annum, of which only USD 4 was spent on public health. There was a significant difference in the MoHFW’s health expenditure at different levels, with the highest healthcare allocation (27%) going to the richest quintile compared to 21% to the poorest quintile (29). Currently, only 2.8% of the country’s GDP is spent on health and over two-thirds of overall health expenditure is paid for privately, out-of-pocket, including for drug purchases and medicines (27,29). Health insurance in Bangladesh practically does not exist, though there have been efforts by NGOs to pilot health insurance schemes (29).

### Planning and Supporting Guidance

Two studies report the improvement in procurement and supply chain processes and monitoring and evaluation capacity during the health sector reform programs (40,42). One study reports that the SWAp era introduced a centralized bulk procurement method for economies of scale. As a result, the procurement process lead time for crucial medical equipment declined from 46 months to 26 months. The percentage of idle procured equipment at health facilities decreased from 57% to 46% (42). The results framework for monitoring progress during the SWAp period improved substantially from 2007 to 2012. Moreover, the health sector reforms introduced two new institutions, the Program Management and Monitoring Unit and the Procurement and Logistics Monitoring Cell, established to reinforce essential program features (42)

### Access to Care

Six studies report on improving the availability of health services for the population and the major barriers to access (20,22,23,29,33,42). One study that compared selected health facility statistics from 1997 to 2011 showed an improvement in the provision of services both in primary (viz. Upazila Health Complexes) and secondary (viz. District Hospital) level facilities during the health sector reform period. In 1981, there were 5350 people for every public hospital bed, which fell to 4293 in 1997 and 3435 in 2010.

Additionally, from 1997 to 2010, hospital beds were up in public hospitals, from 29,106 to 43,996 (i.e., 51%), more than double the growth rate of the total population over time (22%). These improvements were also observed in maternal health services utilization, as better health professionals, drugs, and equipment availability were ensured. Reform initiatives such as revitalising CCs increased the number of service recipients (mainly women and children) over time, from 12 to 38 people per CC per day between 2009 and 2013 (22,33,42). Currently, Bangladesh has a total of roughly 18,000 CCs (22,23).

Despite these improvements, the unavailability of drugs, medical supplies, and family planning products is a constant challenge in many health facilities. One study reports that about 65% of ambulances and other equipment, such as X-ray machines and incubators, are non-functional due to insufficient funds for proper repair or replacement. Essential medications and family planning materials for patients are often hoarded and sold to private vendors. The author explains that these challenges are primarily a result of ineffective supply chain management, insufficient funds, and improper management of available funds (29).

Furthermore, access to care is primarily impacted by maldistribution of available HR in both the private and public sectors, especially between rural and urban areas (with only 16% of physicians vs. 62% of the population in rural areas), the existence of vacant positions (41% of rural-based physician positions left vacant), absenteeism, and the difficult-to-reach location of health facilities (20).

Three studies report on out-of-pocket spending on health services and the impact of health sector reforms on health service affordability (20,22,42). Out-of-pocket healthcare spending currently accounts for 67% of total healthcare expenditure. One of the key NHP objectives is to ensure the availability and affordability of essential drugs through the price control system. Yet, the cost of medications appears to be the most significant component (about 65%) of out-of-pocket costs in Bangladesh. One study explains that the high out-of-pocket spending on medications indicates that Bangladesh’s price-control mechanism for essential drugs is ineffective (22).

Another study also mentions the disparities in spending that either directly or indirectly limits access to health care. Contrary to the policy objective of ensuring that the poor and the disadvantaged have access to health services, public health expenses are favourable for the rich and urban populations. MoHFW spending at and below the upazila level, the level of services largely used by the poor, decreased from 51% in 2003-2004 to 42% in 2005-2006, while the funding for tertiary hospitals and the administration of the Ministry increased. The rich and prominent have more access to these tertiary hospitals, both public and private (20).

Notwithstanding, another study in our analysis reports how health sector reforms led the MoHFW to launch a voucher plan to stimulate demand for basic health services such as delivery, allowing poor pregnant women to purchase maternal health services under a demand-side financing (DSF) modality. Initially, DSF was piloted in 21 upazilas (out of a total 488 upazilas) and then progressively enlarged to 53 during HNPSP. In the pilot locations, DSF successfully improved skilled delivery, boosted safe motherhood practices, and increased facility delivery compared to non-DSF locations (42).

## DISCUSSION

This scoping review aimed to explore the literature on health sector reforms implemented over the past three decades and their impact on the Bangladesh health system. The most significant reform has been the shift from a project-based financing approach to the SWAp, which implemented three reform initiative programs with only a few setbacks. In this sense, Bangladesh is following a global trend (44). In addition to national reform initiatives, the country must also consider and integrate global objectives linked to UHC, Global Health Security (GHS) and Health Promotion. However, the scope of these different programs and their links still need to be clarified to decision-makers, resulting in poor coordination between the various donors. The recent COVID-19 pandemic demonstrated that, during health crises, the synergy between these programs is challenging to achieve, and the emphasis is always on GHS through strengthening health systems to adapt and absorb shocks (5).

Substantial evidence was retrieved for specific health system dimensions such as health sector management, workforce, finance, and planning and supporting guidance. There has been an increase in qualified health professionals. However, many are concentrated in the urban areas, leaving the rural areas in need of attention. These findings mirror those reported by the MoHFW and WHO. Human resources issues in the health sector is a global challenge. However, Bangladesh stands out because of the dual problem of the emigration of its health professionals to wealthier nations and the scarcity of medical personnel in its rural and isolated region. Our review also highlights the high percentage of out-of-pocket payments with no robust insurance system. In terms of access to care, there needed to be more evidence. Reform initiatives such as the revitalization of existing CCs and voucher schemes for pregnant women have improved local rural service utilisation. There has also been an increase in hospital facilities, but the coverage still needs to be improved. The lack of rural health professionals, insufficient supplies and equipment maintenance funds, and decreased public financing for rural health facilities are all noted as barriers to access to care.

### Context-Specific Policy Reforms: the question of ownership

The dependent relationship between the GoB and international donors has led to a strained relationship regarding ownership, also widely found in Africa (7). When visions are aligned, these issues do not come to the forefront. However, in the case of the merger of the health and family welfare sectors of the MoHFW, ownership became a topic of great interest, as in Mali in West Africa where tensions between the two ministries are age-old (45). On the one hand, refusal of this suggested reform could be viewed as a successful demonstration of government ownership, but on the other hand, some DPs saw this as a sign that their time and money were being wasted (46,47). The continuing influence of DPs in low- and middle-income countries (LMICs) has been viewed through different lenses.

Although many of their proposed policy reforms aim to improve health systems and healthcare in the target countries, they can also lead to unintended shocks. As health policy and public health literature mention, decision-makers focus on the content of reforms while neglecting other vital elements such as context, process, and actors (33,42,48). Getting back to the heart of implementation science in the health field is essential to better account for the evidence and reduce the implementation gap. In the case of Bangladesh, DPs should have recognised these elements in their push for policy reforms, as is often the case with travelling model reforms (49,50). In the context of health system resilience, we can also see how policy reforms can result in shocks or disturbances that impact the health system. More and more empirical and conceptual studies are showing how important it is to take account of reforms and events that cause shocks to healthcare systems, in addition to pandemics and other external factors (51–53). In Mali, for example, the lack of health workforce reform, in addition to security challenges, has limited the resilience of the health system (54). As Bangladesh expands its economic output, the relationship between DPs and the national health system will continue to be re-examined.

### Absence of a well-balanced workforce

It is well known that the adequate availability and distribution of qualified health professionals is necessary for health system resilience (54–56). However, Bangladesh’s health workforce shortage and maldistribution show the vulnerability of its health system. One of the most pronounced shortages is for nursing professionals. One of the studies (43) reported prejudice in the way the nursing profession is viewed. Some of this prejudice stems from cultural and religious factors. For example, Hindus may consider interacting with bodily excretions to be a job for the lower castes, while Muslims may be uncomfortable interacting with the opposite gender (57). Cultural issues are rarely considered in health reforms, and examples from Asia and Africa show how these relationships can undermine the quality of care for women (58,59).

The other primary source of prejudice stems from education and professional competence. The capacity of nurses to carry out clinical activities independently without a doctor’s supervision is questioned. Such bias can influence one’s choice to study nursing. Addressing systemic issues in nursing competence requires industry-wide development. The quality of nursing (private) education must be enhanced, and regulations of the profession must be followed through (60). There has been a historical lack of interest in establishing nursing institutions in the country, which has created a bottleneck in the education system and the establishment of career progression routes (57). There is also a low retention of health professionals in rural areas (36,42). Poor working conditions and rampant absenteeism, which are especially common in rural areas, must be addressed to improve the retention of nurses and other health professionals (30,39,60).

### Low Funding for healthcare services

Our review confirms the importance of finance in health care delivery and access to care. Although increased spending does not have a one-to-one ratio with better health outcomes or service delivery, health systems still require a well-funded environment to operate smoothly (61). In Bangladesh, low public funding is prevalent, which leads to high out-of-pocket spending among the population. Adopting structural adjustment programs in Bangladesh could explain this low public financing but also the persistence, as elsewhere in the world, of a neoliberal approach to healthcare systems as proposed by numerous international financial institutions (62,63). There have been several controversies on the negative impact of these programs in the health sector as processes such as the liberalisation of the economy and the privatisation of the private sector cause a reduction in public health financing (23). As elsewhere in the world, attempts at performance-based financing have yet to be very effective, and even the World Bank is now proposing to abandon this system (64). A key challenge is the government’s reliance on DPs and private sector funding to drive health sector reform initiatives while spending more in other sectors. Ultimately, services must be paid for out-of-pocket, putting many at risk of being forced into poverty (6). It is worth noting that the GoB is working to improve finance mechanisms and provide financial protection against high expenditures through its Health Care Financing Strategy 2012-2032 (65). Evaluating its implementation and effectiveness will undoubtedly be very useful.

### Limitations

This is the first comprehensive scoping review of the literature on the impact of health sector reforms in Bangladesh. This study used literature available through the selected databases, so it may have excluded documents, mainly grey literature and documents written in Bangla that may have been available through country-specific organisations. The initial title and abstract screening were conducted by one reviewer in 2021, which may have introduced bias or exclusion of relevant articles. However, we have updated this scope based on articles published up to June 2023, which reinforces the scope of the review. Scoping reviews only provide a descriptive analysis. Many of the included reports, and policy documents did not use defined qualitative or quantitative methods. Thus, one of the limitations of the review is the need for more data to enable an evaluative judgement to be made in the causal analysis of healthcare reforms, which is complex and not possible with this type of scoping review. The next step is data permitting to conduct such a causal analysis using more appropriate methods such as modelling or meta-analysis, which was done to show the effects of user fee exemption policies (66,67). More experimental methods could also be used, but they are rare in this field, as decision-makers often need to remember the impact assessment issues when formulating their public policies (68). In addition, we have noted that the issue of quality of care has received little attention in analyses of reforms, except in a recent scoping review on universal health coverage (69). Lastly, the concept of access to care should have been included in the search strategy as it produced numerous non-relevant results.

## CONCLUSION

Health policy in Bangladesh, as has in many LMICs, historically depended on the guidance of international donor agencies. So there are many sustainability and ownership issues (7), as well as adapting often exogenous reform models to national contexts (49). In addition, power issues in healthcare reforms are at the heart of the process (70,71), including when it comes to the resilience of healthcare systems (53). The role of ideology and power, for example, has been central to the emergence of national health insurance policies in Rwanda (72) and India (73), with some success in terms of effectiveness. Existing reforms still need to address the issue of centralisation and the shortage of formally qualified health professionals such as nurses and technologists. However, health sector reforms have improved some health system functions and access to care despite the abovementioned constraints. There remains room for deeper causal meta-analyses on the specific factors discussed. Lastly, this scoping review has identified the vulnerabilities in the health system that need to be addressed immediately to influence policies that can build a system that is more resilient to emerging stressors such as climate change and epidemics (5).

## Data Availability

All the data used are in the text of this article.

https://www.example.com

## ACKNOWLEDGMENTS

We thank Professor Syed Massud Ahmed for carefully reviewing the article and his constructive comments. We would also like to thank the EHESP School of Public Health Librarians, Sarah Louart for their support with research strategies and article selection (with Matin Mowtushi) and Bertrand Lefebvre’s support to Treasure as part of her internship.

