## Supplementary material for "Health Systems Reforms in Bangladesh: An Analysis of the Last Three Decades": Supp 2

| Database | |
| --- | --- |
| Database | MEDLINE |
| Interface | PubMed |
| First research date | 24 March 2021 |
| Second Research date | 04 July 2023 |
| Filters | Year: 1991 – 2023  Language: English, French |

| Syntax | |
| --- | --- |
| [MeSH Terms] | Medical Subject Heading |
| OR, AND | Boolean operators |
| * | Truncation |

Search strategy

((((((reform*[Title/Abstract]) OR (polic*[Title/Abstract])) OR (health care reform[MeSH Terms])) OR (public policy[MeSH Terms])) OR (health policy[MeSH Terms])) AND ((((("health"[Title/Abstract]) OR ("healthcare"[Title/Abstract])) OR ("healthcare sector"[Title/Abstract])) OR (healthcare system[MeSH Terms])) OR (healthcare sector[MeSH Terms]))) AND ((Bangladesh[Title/Abstract]) OR (Bangladesh[MeSH Terms]))

Total : 1555 articles

| Database | |
| --- | --- |
| Database | SCOPUS |
| Interface | Elsevier |
| First research date | 17 March 2021 |
| Second Research date | 05 July 2023 |
| Filters | Language: English, French |

| Syntax | |
| --- | --- |
| OR, AND | Boolean operators |
| * | Truncation |
| TITLE-ABS-KEY | Title, Abstract and Keywords |

Search strategy

TITLE-ABS ( "reform*" ) OR TITLE-ABS ( "polic*" ) OR INDEXTERMS ( "health care reform" ) OR INDEXTERMS ( "public policy" ) OR INDEXTERMS ( "health policy" ) AND TITLE-ABS ( "health" ) OR TITLE-ABS ( "healthcare" ) OR TITLE-ABS ( "healthcare sector" ) OR INDEXTERMS ( "healthcare system" ) OR INDEXTERMS ( "healthcare sector" ) AND TITLE-ABS ( "Bangladesh" ) OR INDEXTERMS ( "Bangladesh" ) AND ( LIMIT-TO ( LANGUAGE , "English" ) OR LIMIT-TO ( LANGUAGE , "French" ) )

Total: 2186 articles

| Database | |
| --- | --- |
| Database | GOOGLE SCHOLAR |
| Interface | Google |
| First research date | 17 March 2021 |
| Second research date | 04 July 2023 |
| Filters | Year: 1991 – 2023 |

| Syntax | |
| --- | --- |
| OR, AND | Boolean operators |
| * | Truncation |

Search strategy

allintitle: bangladesh health reform OR reforms OR reforming OR policy OR policies

Total: 166 articles (with citations)

| Database | |
| --- | --- |
| Database | WEB OF SCIENCE |
| Interface | Clarivate |
| Research date | 18 March 2021 |
| Second research date | 04 July 2023 |
| Filters | Year: 1991 – 2023 |

| Syntax | |
| --- | --- |
| OR, AND | Boolean operators |
| * | Truncation |
| TS | Title |

Search strategy

TS=(reform* OR polic* OR health care reform OR health policy) AND TS=(healthcare sector OR healthcare system) AND TS=(Bangladesh)

Total : 116 articles
